# Longitudinal Study on Seroprevalence and Immune Response to SARS-CoV-2 in a Population of Food and Retail Workers Through Transformation of ELISA Datasets

**DOI:** 10.1101/2024.01.27.24301877

**Authors:** Abdelhadi Djaïleb, Megan-Faye Parker, Étienne Lavallée, Matthew Stuible, Yves Durocher, Mathieu Thériault, Kim Santerre, Caroline Gilbert, Denis Boudreau, Mariana Baz, Jean-Francois Masson, Marc-André Langlois, Sylvie Trottier, Daniela Quaglia, Joelle N. Pelletier

## Abstract

Since the onset of the global pandemic caused by the emergence and spread of SARS-CoV-2 in early 2020, numerous studies have been conducted worldwide to understand our immune response to the virus. This study investigates the humoral response elicited by vaccination and by SARS-CoV-2 infection in the poorly studied food and retail workers in the Québec City area. The 1.5-year study period spans from early 2021, when vaccination became available in this region, to mid-2022, following waves of virulence due to the emergence of the first Omicron variants. Cross-correlated with data on workplace protective measures, pre-existing conditions, activities and other potentially relevant factors, this longitudinal study applies recently developed ELISA data transformation to our dataset to obtain normal distribution. This unlocked the possibility to use the ANOVA-Welsh method for statistical analysis to obtain a statistical perspective of the serological response. Our work allows the identification of factors contributing to statistically relevant differences in the humoral response of the cohort and strengthens the utility of the use of decentralized approaches to serological analysis.

## Introduction

Following the emergence of the severe acute respiratory syndrome coronavirus 2 (SARS-CoV-2) in Wuhan (China) in late 2019, the World Health Organization (WHO) declared global pandemic status on March 11^th^ 2020 [1–3]. Over the first three years of the pandemic, over 1.3 million confirmed cases and more than 19,000 deaths were documented in the province of Québec alone (current population 8.6 million) [4]. Variants of the virus have appeared and continue to appear worldwide, often triggering new waves of infection [5]. Since December 2020, a growing number of the emerging variants have been classified as “variants of concern” (VOC) by the WHO for being highly contagious, capable of evading post-infection immunity, and by causing a higher number of hospitalizations [5].

Higher antibody detection in infected or vaccinated individuals has been correlated with increased protection efficiency [6–12]. This made serosurveys a central tool for evaluating and predicting the protection level against viral infections within various populations deemed at risk [7]. Studies have been predominantly carried out on cohorts of healthcare workers [13–17], immunocompromised individuals having various pre-existing conditions [18–20], hospitalized COVID-19 patients [13,21–23], the elderly [24,25] and younger individuals [26–29]. Of interest, studies also reported on a higher risk of SARS-COV-2 infection in food and retail workers during the early period (prior to vaccination) [30–34]. These workers were considered at greater risk of infection due to the customer-facing nature of their occupation, which was classified as essential by the Public Health Agency of Canada, even during periods of confinement in Québec [35].

To investigate this population, we performed a study on a cohort of 304 food and retail workers that covered key periods of the pandemic, including the initial vaccination campaigns and the emergence of the Omicron variant. Samples collected in this work were first analyzed via a centralized laboratory platform using a standardized and automated chemiluminescent ELISA assay (hereafter referred to as the “centralized assay”) at the Serology and Diagnostics Facility of the University of Ottawa, which operated in partnership with the Public Health Agency of Canada for COVID-19 serological assays [36]. This platform performed YES/NO determination of SARS-CoV-2 vaccine seroconversion and/or infection events by measuring the IgG antibodies targeting the Wuhan-Hu-1 ancestral strain spike trimeric ectodomain (hereafter referred to as the “spike protein”), the receptor binding domain (RBD) and the nucleocapsid protein [36].

To greatly expand the information held in the samples, we then used a semi-automated in-house colorimetric ELISA assay developed at our decentralized site at the Université de Montréal (hereafter referred to as the “in-house assay”) [37]. With this method, we performed YES/NO detection similar to those of the centralized method to validate coherence of the analyses, using the SARS-CoV-2 Wuhan-Hu-1 ancestral spike protein and nucleocapsid protein. We then broadened the study to determine the cross-reactivity of IgG directed against the spike protein of the SARS-CoV-2 Delta and Omicron VOCs, and assessed the IgM directed against the Wuhan-Hu-1 ancestral spike and nucleocapsid proteins. Importantly, integration of a recent mathematical model [38] allowed transformation of the dataset for normalization, enabling calculation of statistical differences in antigen levels between vaccinated and unvaccinated individuals and between individuals having different job categories or potential risk factors.

This work is part of a collective effort and stresses the importance of rapidly developing adaptable, decentralized tests for population-level immune surveillance in response to a pandemic, even before centralized testing is available [37,39]. To our knowledge, no other study has reported such an extensive 18-month longitudinal investigation during the key periods preceding and following the emergence of Omicron in a cohort of highly vaccinated food and retail workers, using both centralized and customized methods to analyze two types of immunoglobulin, three types of epitope, and antigens to three VOC [40,41], [42]. This study will inform strategies and measures to be implemented in the event of a future pandemic.

## Materials and Methods

### 1. Cohort composition and sample collection

All recruitment, data collection interviews and sample collection activities were carried out at the Centre Hospitalier Universitaire (CHU) de Québec-Université Laval and have been described in detail [39]. Briefly, the cohort included 304 food and retail workers from the Capitale Nationale (76%) and Chaudière-Appalaches (24%) administrative regions of the Canadian province of Quebec. Individuals provided written informed consent (approved by the “Comité d’éthique de la recherche du CHU de Québec-Université Laval”, registration number 2021-5744). The Canadian Immunity Task Force (CITF) questionnaire [60] was used for data collection and data collected during interviews have been reported [39]. With an average age of 41.3 years, 58 % were female and 42% were male; nearly half (49%) worked in restaurants or bars, 37% in grocery stores and 14% in hardware stores. None had a history of hospitalization linked to SARS-CoV-2 [39].

The observation period spanned 1.5 years, from April 2021 to October 2022. A total of 121 participants reported at least one positive test during the study period. Participants attended five evenly spaced visits between April 2021 and October 2022. A blood sample and information on SARS-CoV-2 risk factors, symptoms, antigen or PCR test results, and vaccination status were collected at every visit. In addition, information regarding potential risk factors (i.e., demographic, socioeconomic, behavioral, clinical, and occupational) was obtained, allowing multifactorial analysis of the cohort.

At each visit, a 30 mL blood sample was collected in 6 mL tubes (BD Vacutainer 367815). Sera collected between June 4^th^ and 9^th^, and between July 2^nd^ and 8^th,^ 2020, from eight individuals (aged between 20-55 years old; 7 females and 1 male) who had never tested positive for SARS-CoV-2 were pooled and used as the negative control. Sample tubes were gently inverted, held at room temperature for 15–30 min to allow clotting and spun at 1600 g for 15 min. Serum samples (1 mL aliquots) were transferred into cryovials (Sarstedt Inc., product 72.694.006), frozen in an upright position at -20°C and stored at -80°C until shipment on dry ice to the assay site, where they were maintained at -80°C until use.

### 2. SARS-CoV-2 viral antigens

The National Research Council of Canada (NRC) provided trimeric recombinant SARS-CoV-2 spike proteins (Wuhan-Hu-1 (SmT1, lot PRO1-429), Delta (SmT1v3 (B.1.617.2)), and Omicron BA.1 (SmT1(BA.1), lot PRO7911-2]), as well as the spike receptor-binding domain (319–541 RBD) produced as described previously.[61,62] The NRC also supplied the SARS-CoV-2 nucleocapsid NCAP-1.[63]

### 3. In-house ELISA (Enzyme-linked Immunosorbent assays)

The semi-automated in-house colorimetric ELISA was performed as described in [45] and [37] with some modifications by adapting a semi-quantitative ELISA protocol from previous work [64–66]. Briefly, 96-well immunoassay plates were coated with 100 μL of the relevant SARS-CoV-2 antigen and incubated overnight at 4 °C. They were then washed and blocked with 100 μL of 3% (w/v) skimmed milk powder for 1h at room temperature (RT) and then washed again. The human serum samples were inactivated by incubating them in a block heater at 56 °C for one hour and diluted as specified (1: 15,000 for anti-spike IgGs, 1: 200 for anti-nucleocapsid IgGs, and 1: 100 for anti-spike and anti-nucleocapsid IgMs). Aliquots (100 μL) of the inactivated, diluted human serum samples were then added in the prepared immunoassay plates and incubated at RT for one hour. The plates were washed, and 100 μL of the appropriately diluted host-specific secondary antibody was added (1: 30,000 Goat anti-human IgG HRP Life Technologies (Invitrogen) Catalog #31413 or 1: 10,000 Goat anti-human IgM (µ-chain specific) HRP Sigma-Aldrich Catalog # A6907), covered to block out light, incubated at RT for 1h, and then washed again. 100 μL of 3,3ʹ,5,5ʹ-tetramethylbenzidine (TMB) was then added and incubated at RT for 1h, followed by the addition of 100 μL of 2 M HCl for color development. Absorbance was measured at 450 nm.

Quality control was performed using a calibration standard to ensure inter-day reproducibility throughout the testing period. The calibration standard consisted of pooled clinical samples from participants who were confirmed COVID-19-positive by PCR test. Independent replicates (n = 45, performed on different assay plates over several months) performed for each antigen and immunoglobulin type determined the standard signal range for each antigen. The standard was included on each ELISA plate (3 wells on each 96-well plate). When the value obtained for the standard fell outside of the acceptable range (+/- 25% of the median value), assay results for that plate were excluded. We note that the median value was determined retrospectively, in that the 45 independent replicates were performed alongside the query samples. According to that median, we then retrospectively evaluated which 96-well plates should be excluded. Those assays were repeated. The COVID-19-negative control (described above) was also included on each ELISA plate.

Independent replicates performed for each specific antigen and immunoglobulin type (n = 45, performed on different assay plates over several months) established the negative control range for each target. Positivity thresholds were established as values above the mean of negative controls for a given target plus one standard deviation. Again, positivity thresholds were determined retrospectively.

### 4. ELISA (Enzyme-linked Immunosorbent assays) at the uOttawa High Throughput Serology and Diagnostic Facility

The automated chemiluminescent ELISA, referred to as the ‘centralized assay’, was described in [36]. Briefly, automated chemiluminescent ELISAs were performed using MicroLab Star robotic liquid handlers (Hamilton, USA) and a 405 TS/LS LHC2 plate washer (Biotek Instruments; all wash steps included four washes with 100 μL PBST). All incubations were done at room temperature with shaking at 500 rpm. Antigens (Wuhan-Hu-1 ancestral spike, 319–541 RBD, and NCAP-1 nucleocapsid) were diluted in PBS and dispensed into the wells of a 384-well high-binding polystyrene Nunc plate (Thermo Fisher Scientific, #460372) at a final amount of 50 ng/well. The plates were centrifuged at 216 × *g* for 1 min to ensure even coating, incubated overnight, rocking at 4°C, and washed. Wells were blocked with 80 μL of 3% w/v skim milk powder dissolved in PBST for 1 h and then washed. Samples and controls were diluted as indicated to a final concentration of 1% w/v skim milk powder in PBST, and 10 μL was added to each well from a 96-well source plate. Plates were incubated for 2 h, and wells were washed. Secondary antibodies (as in the colorimetric assay) were diluted as indicated in 1% w/v skim milk powder in PBST, and 10 μL was added to each well. After incubation for 1 h, the wells were washed, and 10 μL of ELISA Pico Chemiluminescent Substrate (Thermo Scientific; diluted 1:2 in MilliQ H_2_O) was dispensed into each well. After a 5 min incubation with shaking, plates were read on a Neo2 plate reader (BioTek Instruments) at 20 ms/well and a read height of 1.0 mm [36].

### 5. Data transformation and statistical analysis

#### Standard curve of the ELISA standard

For calibration purposes, a parent solution referred to as the “standard” and consisting of a pool of 13 COVID-19-positive samples, was created and used throughout the study. To model the relationship between absorbance and concentration, a 12-step serial dilution of the standard was prepared. As the actual antibody concentration of the standard was unknown, the most diluted sample was arbitrarily set at a concentration of 1. Each subsequent sample was assigned a relative concentration twice that of the previous sample. Consequently, the serial dilutions included relative concentrations (R_c_) of 1, 2, 4… to 2048. ELISA of the serial dilutions was performed against each relevant antigen, in quadruplicate on each of three separate plates for a total of 12 replicates of each dilution. Each serial dilution plate also contained a negative control (pool of COVID-negative samples) and a blank control (buffer only).

#### Modeling the relationship between absorbance and concentration

For each antigen, the mean ELISA absorbance of each dilution of the standard was then related to the dual log of its relative concentration (R_c_) to plot a nonlinear calibration curve of O.D. versus log_2_[R_c_]. To describe this relationship, we used a five-parameter logistic regression model [38]:

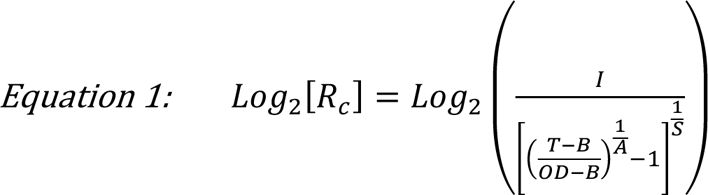

Two of the 5 model parameters (B and T) were fixed (B = median of blank controls, T = technical limit of the spectrophotometer). The three further parameters (A = curve skewness, I = R_c_ at the inflection point, I = R_c_ at the inflection point if A = 1) were predicted using the R script published in [38]. The script uses the Levenberg-Marquardt algorithm and the nlsLM function from R’s minpack.lm package [38].

#### Quality control of the model for each antigen

To confirm the robustness of this method, we reintroduced the serial dilution data (O.D.) from the ELISA for each antigen into the equation using the parameters predicted by the model (again using the code available in [38]) to compare the trend of the experimental data points to the model, confirming the accuracy of each model.

#### Data transformation and analysis

The experimental ELISA data were fed into their respective antigen model for transformation. The limits of these models are their asymptotes. Any data point below the bottom asymptote could not be transformed and was removed from statistical analysis. There were no data points above the top asymptote. The number of data points removed for each antigen dataset is available in **Table S1**.

## Results and Discussion

### An overview of seropositivity and IgG levels in food service and retail workers from April 2021 to October 2022

Higher binding antibody titer in infected or vaccinated individuals has been correlated with increased protection efficiency [6–12], allowing the evaluation of a population’s protection level and predicting the longevity of protection [7]. During the 1.5-year study period, blood samples from 304 food and retail workers were collected at five visits, each spanning a period of 4 to 7 months (**Fig. 1**). In total, 1299 samples were collected. The cohort composition is summarized under Methods, and complete details of the recruitment method and data collected by interview have been reported [39]. The percentage of fully vaccinated participants (defined by the Canadian government as having received at least two doses of one or a combination of approved vaccines [43]) was already 48% by the first visit and reached 97% at visit 5 (**Table S2**). Samples collected during the study period were analyzed with the following objectives: (i) to determine the trend of humoral immunological response over time and in response to critical pandemic events, such as the emergence of new variants; (ii) to identify any relevant statistical differences related to the workers’ occupations, their vaccination status, protective measures at their workplace as well as their underlying health conditions and lifestyle.

**Figure 1.**
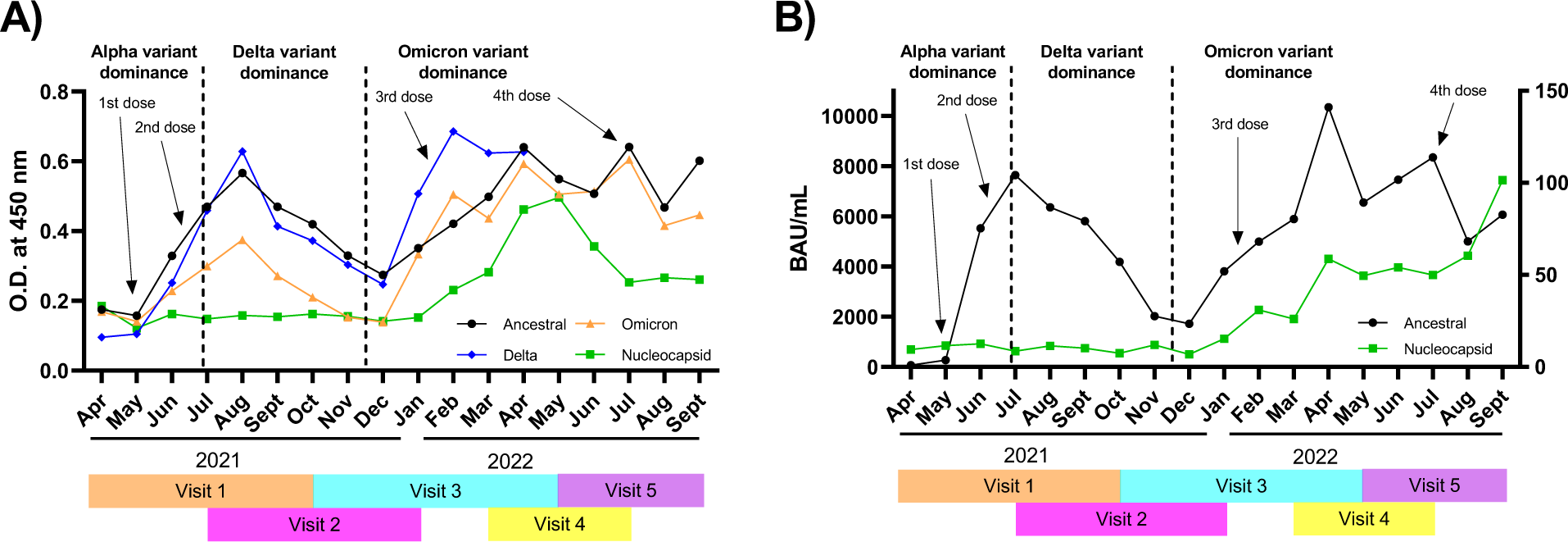
Overview of ELISA from all samples assayed over 18 months (n = 1290). **A)** In-house ELISA, reporting the mean monthly O.D._450_. IgG levels for the ancestral, delta and Omicron spike protein variants and the nucleocapsid were measured (see colored legend). Positivity threshold: ancestral spike = 0.031; delta spike = 0.033; Omicron spike = 0.040; nucleocapsid = 0.048. **B)** Centralized ELISA, reporting the mean antibody units per mL (BAU mL^-1^). IgG levels for the ancestral spike protein and the nucleocapsid were measured (see colored legend). Visits at which samples were taken are indicated in different colors under each graph. The approximate availability of vaccine doses to the general population in the province of Québec is indicated by arrows [47]. Note the scales on the y-axes of (**B**): left axis for ancestral spike, right axis for nucleocapsid.

It is advisable to use the spike protein as a reference in population-based seroprevalence studies, as the humoral immune responses it elicits are strong [44] and it is the antigen used for nearly all approved COVID-19 vaccines. IgG and IgM levels specific to the spike protein ectodomain, as well as the SARS-CoV-2 nucleocapsid protein, were determined for the ancestral strain and the Delta and Omicron VOCs, using a previously established in-house ELISA [37] [45]. The use of a calibration standard made up of pooled clinical samples from PCR-diagnosed COVID-positive participants ensured reproducibility. The standard was included on each ELISA plate, enabling the retrospective establishment of a standard signal range (median +/- 25%) for each antigen. Only plates where the standard signal was within this range were considered valid; invalid assays were repeated as needed. A pool of eight pre-COVID-19 serum samples was used as a negative control and was tested with each specific antigen and immunoglobulin type.

Our objective was first to provide a YES/NO answer to SARS-CoV-2 vaccine seroconversion or infection and then to report on statistical differences in antibody response levels between populations of workers in our cohort. Positivity thresholds (YES) were established as values above the mean of negative controls for a given target plus one standard deviation. The positivity thresholds were determined retrospectively after most samples had been tested (see Methods). The ELISA data were either used as is (YES/NO response) or further transformed into relative concentrations (R_c_) by fitting them to a logistic function obtained using a calibration curve for each antigen for statistical analysis. [38]

Validation of the YES/NO results obtained with the in-house ELISA was achieved by analyzing the same clinical samples at the centralized testing site, using antigens from the same source. Seropositivity was determined at the centralized testing site by ELISA to IgG against both the ancestral Wuhan-Hu-1 strain spike protein ectodomain and receptor-binding domain (RBD) and against the nucleocapsid protein [24,36]. A World Health Organization (WHO) reference standard was used to calibrate the assay [36]. The scaled luminescence values were measured and converted to titer in the form of antibody units per mL [24].

For statistical analysis, calibration curves were obtained by serial dilution of the standard [37,38]. As the standard solution was of unknown concentration, the most diluted sample was arbitrarily set at a concentration of 1, hence the term “relative concentration”. This transformation made it possible to normalize and statistically analyze the data using Welch’s ANOVA, which is only applicable to normally distributed data, yet allows for variance inequality (see Methods) [38].

### Seroprevalence in the population of workers over the timeline of the study

The monthly means of anti-spike (ancestral Wuhan-Hu-I strain) and anti-nucleocapsid IgG levels obtained with the in-house ELISA (**Fig. 1A**, reported as mean O.D. values) were compared with the centralized ELISA (**Fig. 1B**, reported as mean BAU/mL). Concordant trends of seroprevalence were observed by both methods for ELISA to the ancestral variant: anti-spike IgG levels peaked around July/August 2021 during Alpha variant dominance, coincident with the 2^nd^ mass vaccination campaign in the region, and in February to July 2022 during Omicron dominance, coincident with the 3^rd^ mass vaccination campaigns in the region; these observations are consistent with the trends observed more broadly in the Canadian population over the same period [46]. Concordant trends with the anti-nucleocapsid IgG levels (**Fig. 1A** and **B**) were also observed: nucleocapsid signal remained low because relatively few people were infected (**Table 1**) until the highly infectious Omicron variant was reported in Québec on November 29^th^, 2021 [5]. The individual ELISA results are provided in **Fig. S1** (in-house ELISA) and **Fig. S2** (centralized ELISA).

**Table 1:** Comparison of vaccine and natural seroprevalence in vaccinated participants between in-house and centralized ELISA.

|  | Visit 1 |  | Visit 2 |  | Visit 3 |  | Visit 4 |  | Visit 5 |  |
| --- | --- | --- | --- | --- | --- | --- | --- | --- | --- | --- |
| ELISA method | In-house | Centralized | In-house | Centralized | In-house | Centralized | In-house | Centralized | In-house | Centralized |
| Total samples | 304 | 304 | 297 | 296 | 291 | 289 | 194 | 198 | 194 | 194 |
| Vaccine Immunity <sup>1</sup> | 83% | 79% | 72% | 93% | 88% | 92% | 93% | 98% | 95% | 99% |
| Natural immunity <sup>2</sup> | 22% | 9% | 10% | 8% | 38% | 16% | 56% | 52% | 42% | 59% |

<sup>1</sup> Defined by positive anti-spike signal for in-house ELISA or the combination of positive anti-spike and anti-RBD signals for the centralized ELISA.
<sup>2</sup> Defined by positive anti-nucleocapsid signal for in-house ELISA and the combination of positive anti-nucleocapsid and anti-RBD signals for the centralized ELISA.

We further investigated seroprevalence in this population by monitoring the IgM directed against the Wuhan-Hu-1 ancestral spike and nucleocapsid using the in-house ELISA assay. The IgM ELISA signals observed using ancestral spike and nucleocapsid antigens were consistently lower than the corresponding IgG signals, as reported in prior studies (**Fig. S3**)[37,48]. The pattern of rapid production and decline of anti-SARS-CoV-2 IgM is consistent with other reports [37,48–50]. This confirms the utility of reporting on IgM to monitor populational seroprevalence shortly after vaccination or infection events, while illustrating the greater overall practicality of reporting on IgG over longer observation periods.

### Seropositivity in the population of workers over the timeline of the study

The ELISA data allowed determination of seropositivity for each individual, at each visit (**Table 1**; **Fig. S1** and **Fig. S2**). In this standard YES/NO analysis, concordant patterns of vaccine seroprevalence (anti-spike IgG, ancestral strain) was observed between the in-house and centralized ELISA methods. A strong correspondence was also observed for naturally acquired seroprevalence (anti-nucleocapsid IgG) (**Table 1**). Furthermore, the anti-nucleocapsid IgG data is fully consistent with infection data, including reports of PCR-positive test or positive self-test, where infection among this cohort was low (fewer than 5 cases per 100 person-years) during the first waves, rapidly increasing with the emergence of Omicron (peaking at 80 cases per 100 person-years during the fifth wave) [39]. These observations reinforce the validity of using in-house ELISA as a reliable instrument for populational studies during the early phases of a pandemic [37].

The scope of the study was broadened by monitoring Delta and Omicron anti-spike IgG levels using the in-house ELISA assay, highlighting the adaptability of this method (**Fig. 1A**). Before December 2021, during the period where serological response resulted predominantly from vaccination using ancestral spike antigen and before the appearance of the Omicron VOC, the IgG ELISA response using the Delta spike antigen was indistinguishable from that using the ancestral Wuhan-Hu-1 spike antigen. This is consistent with prior reports of effective cross-reactivity between these strains [37,51,52]. As the study progressed, the Omicron variant became dominant. In contrast with the Delta variant, and also consistent with a prior report, cross-reactivity with the Omicron spike antigen was weaker but nonetheless consistent with immune protection even as the viral strains continued to evolve (**Fig. 1A** and **1B**) [53].

### Statistical correlation between vaccination and antigen levels

COVID-19 vaccines were made available to the general adult population in Québec beginning in March 2021 [47], coinciding with the beginning of this study. The Astra Zeneca COVISHIELD (Vaxzevria) was rapidly followed by the Pfizer-BioNTech and Moderna Spikevax RNA-based vaccines; all vaccines initially approved for use in the Québec region were based on the ancestral spike antigen.

Over the course of this 1.5-year longitudinal study, vaccination status increased even as long-term participation in the study decreased: at the first visit (V1), 48% (146 of 304 the study participants) were fully vaccinated (had received at least two doses of one or a combination of the approved vaccines [43]), V3 saw 65% (188/291 participants) fully vaccinated and V5 saw 97% (188/194) fully vaccinated (**Table S2**). We note that the increase in vaccination status from 65% at V3 to 97% at V5 resulted entirely from the drop from 291 to 194 study participants. Interestingly, all who withdrew from the study between V3 and V5 were non-vaccinated. These numbers reveal that vaccination status was strongly correlated with long-term engagement in the study and caution against assigning a significance of the near-perfect vaccination status at V5 to any criterion investigated here.

The in-house ELISA datasets were statistically analyzed to determine correlations of antibody levels with vaccination. Specifically, ELISA reporting on IgG using the Wuhan-Hu-1 ancestral spike, Omicron spike and nucleocapsid antigens were analyzed to determine the correlation between vaccination events and seroconversion, and the impact of viral evolution to the Omicron VOC on cross-reactivity. We note that, following seroconversion for anti-nucleocapsid IgG in any individual, further samples from that individual were excluded to eliminate the impact of infection on the immune response.

We performed statistical analysis of the in-house ELISA dataset by Welch’s ANOVA test. First, mathematical data curation and transformation was performed, fulfilling the requirement that datasets adopt a normal distribution; the unequal variances of the resulting dataset precluded application of ANOVA and indicated Welch’s ANOVA as appropriate. Then, the relationship between ELISA absorbance and relative IgG concentration was modeled, using a COVID-19-positive standard and applying a regression model, as previously reported by others; the data fit a five-parameter logistic function, detailed in Methods [38]. Comparison of ELISA standard curves to the model for each antigen shows excellent concordance (**Fig. 2**). Having confirmed the accuracy of each model, the in-house ELISA datasets were transformed using their respective antigen model and statistically analyzed by Welch’s ANOVA test.

**Figure 2.**
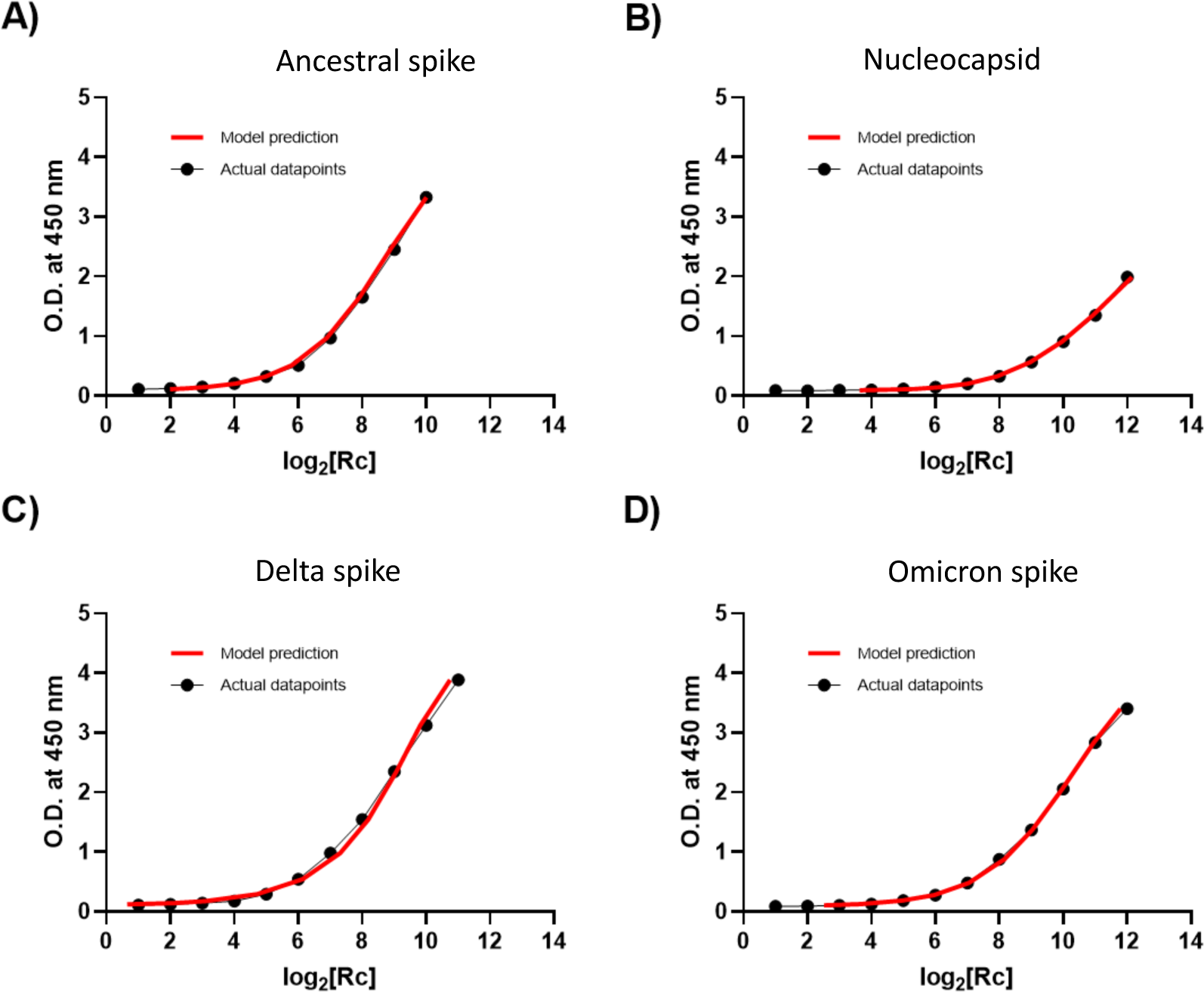
Comparison of serial dilution ELISA data with model prediction. ELISA data for IgG were compared with model predictions for **A)** ancestral spike, **B)** nucleocapsid, **C)** delta spike and **D)** Omicron spike antigens. X-axis: dual logarithm of the relative concentration of serial dilutions (experimental data points in black, model predicted points in red). Y-axes: O.D. of serial dilutions.

We observed clear statistical differences in anti-spike IgG levels between unvaccinated individuals and those who have received a greater number of doses, up to a maximum of four doses (**Fig. 3**). Analysis of cross-reactivity with the Omicron variant spike antigen gave similar results, although the statistical significance was generally greater. When the analysis was repeated with inclusion of samples following seroconversion for anti-nucleocapsid IgG, the trend was conserved with an increase statistical difference (**Fig. S4**). Although this may be due to the greater number of datapoints analyzed, it is more likely due to the additional immunity-provoking event of the infection in the nucleocapsid-seroconverted individuals. In addition to providing an additional event of antigen exposure, vaccination and infection together have been shown to provide a greater response than either alone [54].

**Figure 3.**
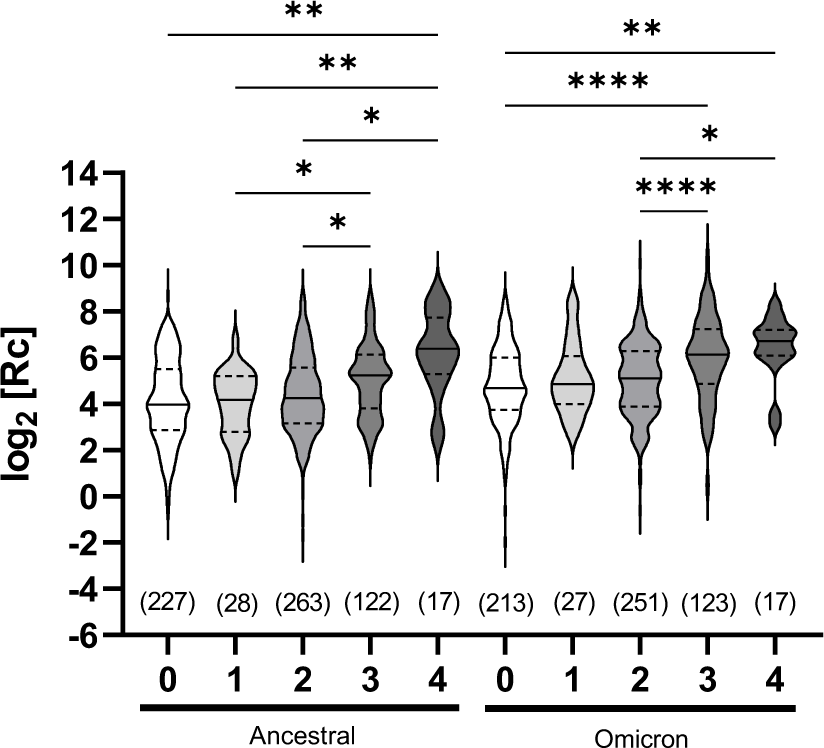
Comparative assessment of humoral immune response in unvaccinated and vaccinated, nucleocapsid-negative individuals. ELISA assays measuring ancestral or Omicron anti-spike IgG levels were analyzed collectively (all visits together). Each grouping includes all data collected (n is given) for that number of vaccine doses (0 to 4, x-axis); ‘n’ excludes samples of individuals following determination of seropositivity for anti-nucleocapsid IgG and includes any sample taken at different visits for the same individual whose vaccination status did not change. X-axis: number of vaccine doses received (minimum 7 days post-vaccination). Y-axis: data points after logarithmic transformation. The median (solid line) and quartiles (dashed lines) are shown in the violin plots; no outliers were removed. Statistical significance: *, p < 0.05; **, p < 0.01; ****, p < 0.0001.

It is important to note that the samples were collected without regard for the dates of vaccination of the individuals. Therefore, the observed serological response is averaged over many months post-vaccination for this population. This is seen in the modest increase (below statistical significance) in signal resulting from two doses of vaccine (‘primovaccination’) compared to non-vaccinated. Since sampling continued for months prior to the 3^rd^ vaccination campaign, it should not be interpreted as poor serological response as it is subject to the expected waning of humoral response over time following primovaccination [55]. A marked increase in median anti-spike IgG levels in samples following 3 or 4 vaccine doses was observed, consistent with other reports [56]. We note that the dataset included few participants having received a single vaccine dose, as many participants had already been vaccinated twice at their first sample collection visit.

Using the same mathematically transformed ELISA dataset, we monitored for any statistical differences in seroprevalence to the spike antigen as a result of the type of vaccine administered. Two vaccine doses (no matter the combination of vaccine products used) resulted in no statistical difference with the unvaccinated individuals (**Fig. 4A**). For three vaccine doses, a statistical difference between unvaccinated individuals and fully vaccinated participants was observed for the combination of three RNA-based vaccines (no matter the combination of Moderna/Pfizer-BioNTech products used); cross-reactivity with the Omicron spike antigen gave a higher statistical significance. Although the trend was maintained for the combination of Astra-Zeneca/2RNA vaccines, it was not statistically significant (**Fig. 4B**). However, upon including the nucleocapsid positive individuals in the analysis (**Fig. S5 A** and **B**), greater statistical differences were seen. As above, this could be due to the greater number of datapoints analyzed, and to the additional immunity-provoking event of the infection in the nucleocapsid-seroconverted individuals.

**Figure 4.**
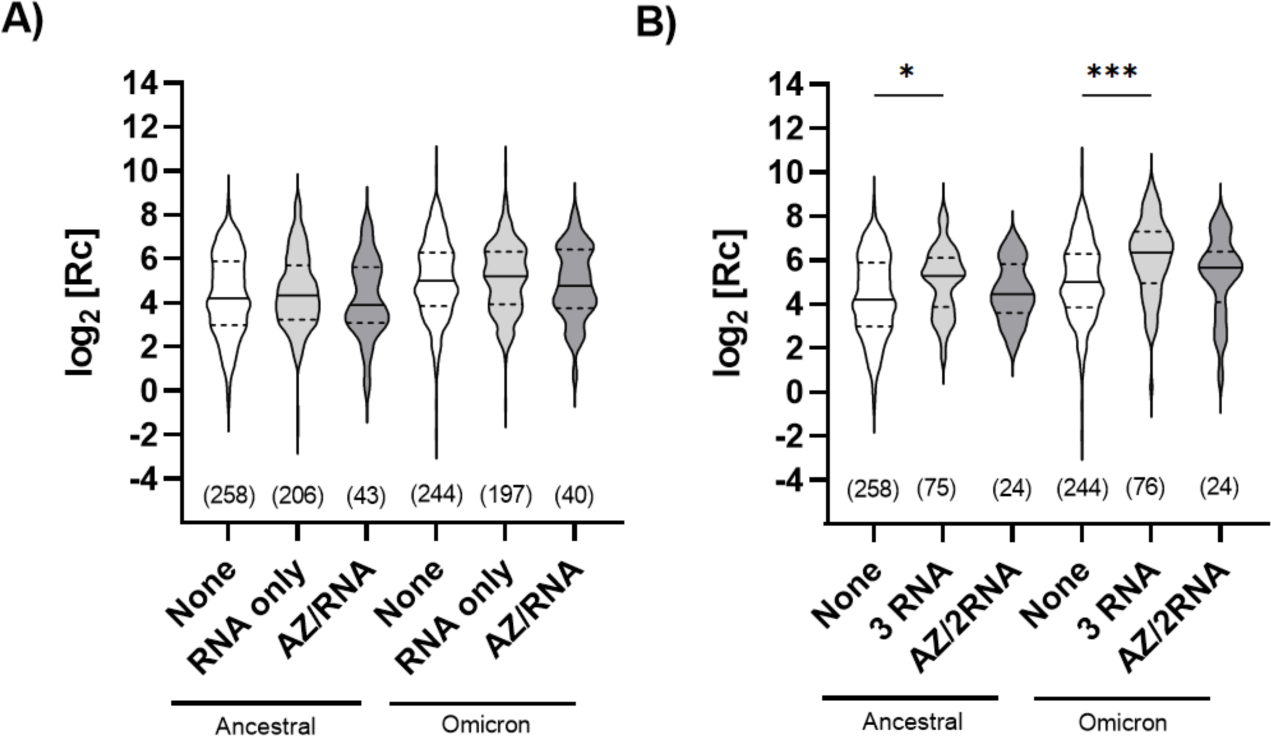
Comparative assessment of humoral immune response for different vaccine types. ELISA assays measuring ancestral or Omicron anti-spike IgG levels were analyzed collectively (all visits together) following administration of **A)** 2 vaccine doses or **B)** 3 vaccine doses. Each grouping includes all data collected (n is given) for that type of vaccine; ‘n’ excludes samples of individuals following determination of seropositivity for anti-nucleocapsid IgG and includes any sample taken at different visits for the same individual whose vaccination status did not change. “RNA only”: either the Pfizer-BioNTech Comirnaty® mRNA COVID-19 vaccine, the Moderna Spikevax® mRNA COVID-19 vaccine, or a combination of both. “AZ/RNA” and “AZ/2RNA”: one dose of the AstraZeneca COVISHIELD® viral vector-based COVID-19 vaccine and one or two doses of an RNA vaccine, respectively. Individuals who had received the AstraZeneca vaccine only, or two doses of AstraZeneca and one of an RNA vaccine, were excluded as there was insufficient data to allow statistical analysis. Y-axes: data points after logarithmic transformation. The median (solid line) and quartiles (dashed lines) are shown in the violin plots; no outliers were removed. Statistical significance: *, p < 0.05; ****, p < 0.0001.

Our findings are consistent with the report that vaccine efficacy is roughly comparable, reaching around 90% seroconversion when the two doses administered are from an mRNA vaccine (i.e. Pfizer and Moderna), or when the first dose administered is AstraZeneca (viral vector vaccine) but followed by a second dose with an mRNA vaccine [57].

### Statistical analysis of antigen levels in relation to the workplace in an under-reported yet highly exposed population

The past years have seen a worldwide, concerted effort to understand the human immunological response to SARS-CoV-2. Our focus on the humoral immunological response of the underrepresented yet highly exposed group of food service and retail workers has yielded a comprehensive dataset spanning the 18-month period during which vaccination was deployed and the virus evolved into several VOCs. We solicited the participation of individuals working in bars, restaurants, grocery stores and hardware stores because these jobs, by their very nature, require direct contact with customers and are therefore considered at greater risk than other types of work. Workers in these sectors in Québec often have below-average incomes and no health-related benefits [39]. A certain variability of risk exists in this cohort: while grocery stores and hardware stores remained open throughout the pandemic in Québec, bars and restaurants were periodically closed by health authorities. However, the use of protective equipment was better enforced in grocery and retail stores than in restaurants and bars, where clients stayed longer and removed masks to consume food and beverages [39].

We observed no overall statistical difference in humoral response between the different occupations according to ELISA when the data from all visits were analyzed collectively (all visits together) (**Fig. 5A** and **B**). However, certain trends emerged upon stratification by visit (**Fig. 5C** and **D**), specifically over the period of the two first visits, during which the cohort mostly became fully vaccinated (**Fig. 1**) but where the rate of infection-acquired immunity was low (**Table 2**). At visit 1, ancestral strain anti-spike IgG levels were statistically higher for the individuals working in restaurants than in any of the three other areas of occupation (**Fig. 5C**); at visit two, anti-spike IgG levels were statistically higher in those working in restaurants and hardware stores. Bars were closed in the region investigated for 3 of the 10 months spanning visit 1, coincident with evening curfews; this measure of health protection appears to have been successful in reducing exposure of bar workers relative to the other areas of occupation as nucleocapsids levels for this group are lower than for the others. In contrast, grocery and hardware stores were open throughout visit 1; restaurants were also open, although they were restricted to limited capacity by periods. Interestingly, while restaurant and hardware store workers had the highest level of anti-nucleocapsid IgG response at visit 1, bar and grocery store workers had higher levels from visit 2 on (although only statistically significant for visit 2 itself) (**Fig. 5D**).

**Figure 5.**
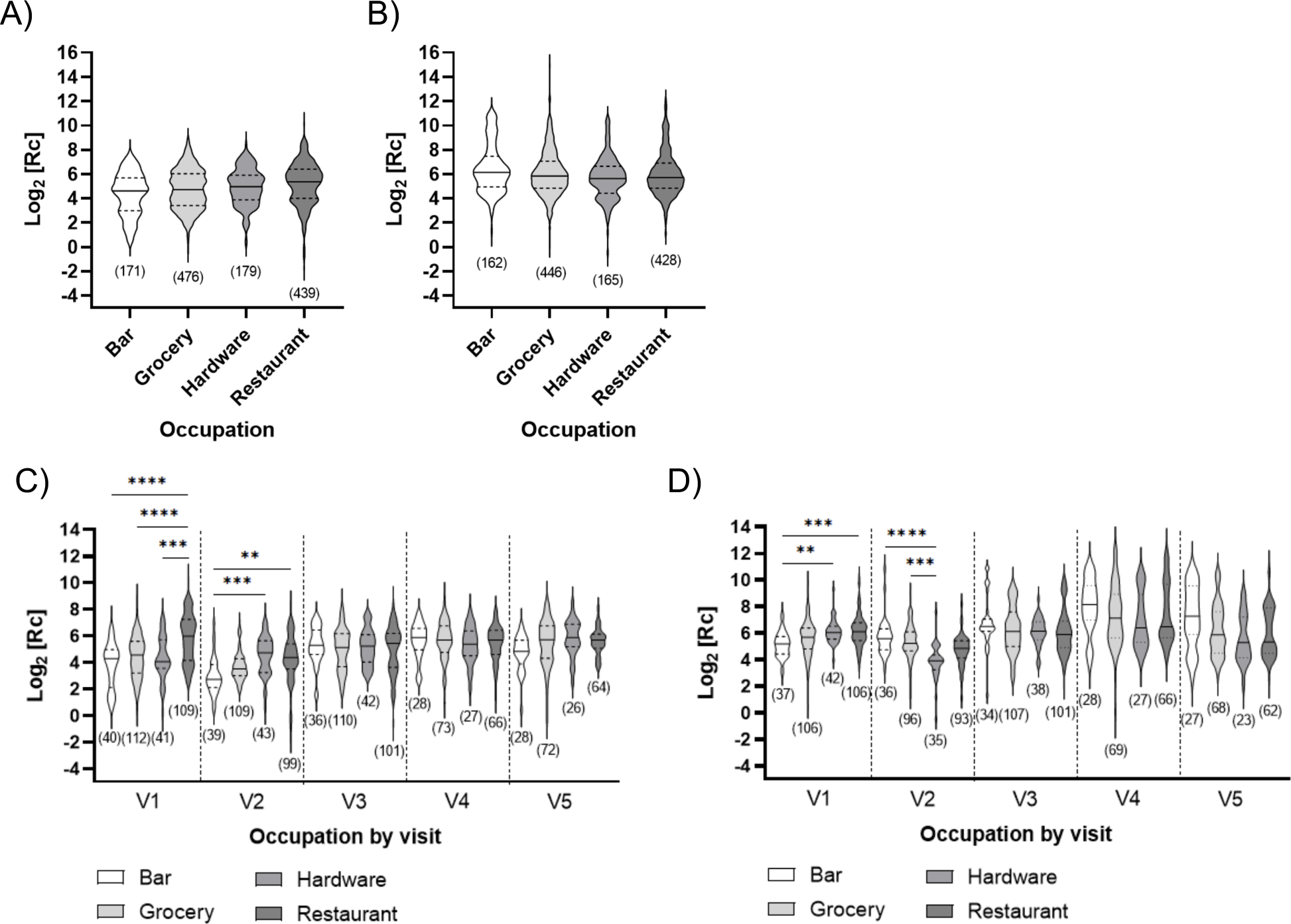
Effect of different occupations on the humoral immune response. ELISA measuring **A)** ancestral anti-spike or **B)** anti-nucleocapsid IgG levels analyzed collectively (all visits together), or **C)** ancestral anti-spike and **D)** anti-nucleocapsid IgG levels stratified by visit (V1 to V5) for serum samples collected during the study period. Results are shown in logarithmic form, post-transformation. The median (solid line) and quartiles (dashed lines) are shown in the violin plots; no outliers were removed. The number of data points (n) per plot is shown. Statistical significance: **, p < 0.01; ***, p < 0.001; ****, p < 0.0001.

An important and contentious subject throughout the pandemic has been whether wearing a mask contributes to protection against exposure to the virus. There is some evidence in the literature that masks provide useful protection [58]. We analyzed self-reported mask-wearing habits at the workplace, finding no statistical difference in the level of anti-nucleocapsid IgG (infection-induced immunity) for all visits, with the exception of visit 4 (**Fig. 6**). Data for visit 4 were collected during the peak of the highly infectious Omicron variant (**Fig. 1)**. Here, more people (65) reported not wearing masks than in previous visits (5 in visit 1, 11 in visit 2 and 21 in visit 3), and their anti-nucleocapsid IgG levels were significantly higher than those of people wearing masks during the same period. We hypothesize that the incidence of the Omicron variant and more relaxed mask-wearing habits contributed to the observed difference.

**Figure 6.**
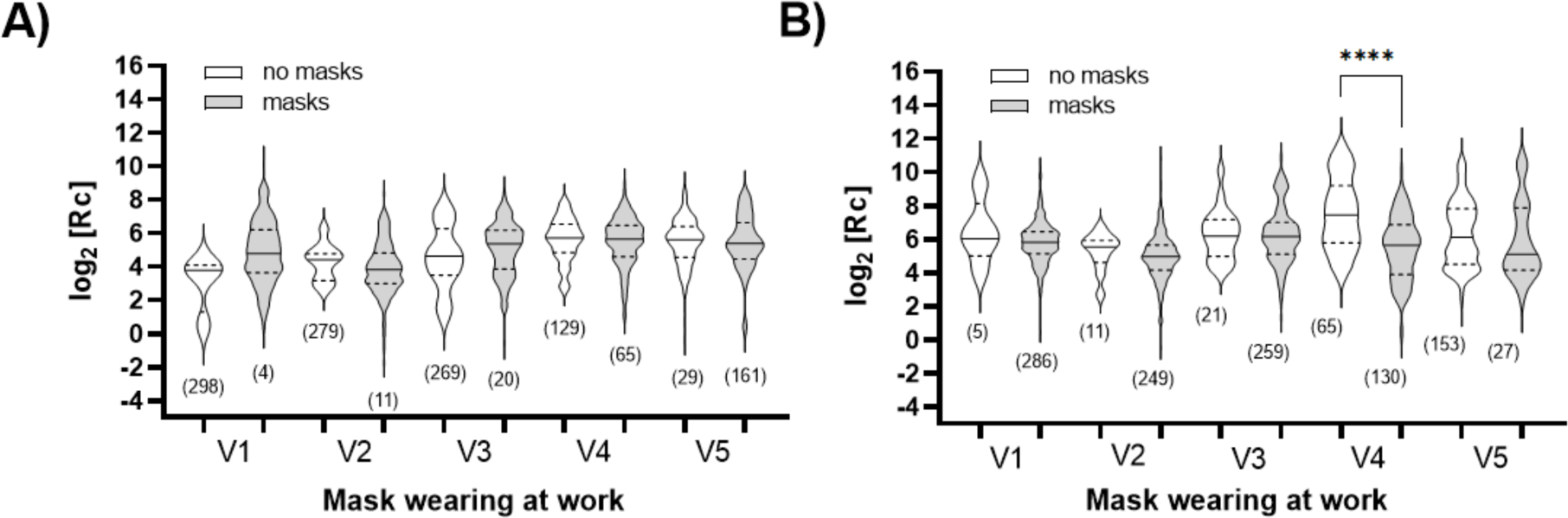
Impact of wearing a mask in the workplace on humoral immune response. ELISA measuring **A)** ancestral anti-spike and **B)** anti-nucleocapsid IgG levels analyzed by visit (V1 to V5). Results are shown in logarithmic form, post-transformation. The median (solid line) and quartiles (dashed lines) are shown in the violin plots; no outliers were removed. The total number of data points (n) per plot is shown. Statistical significance: ****, p < 0.0001.

We found no significant differences for participants who used public or private transport to get to and from the workplace (**Fig. S6**) or for those who reported being in contact with infected people (**Fig. S7**).

### Statistical findings unrelated to work-related activities

While the main focus of this study was to analyze the effects of vaccination and exposure for this cohort, we had the opportunity to examine other factors thanks to the information provided by participants during the survey at each visit, including level of education (**Fig. 7A**), chronic illnesses (**Fig. 7B**), age (**Fig. 7C**), body mass index (BMI) (**Fig. 7D** and **Fig. S8**) and participants’ smoking habits (**Fig. 7E**).

**Figure 7.**
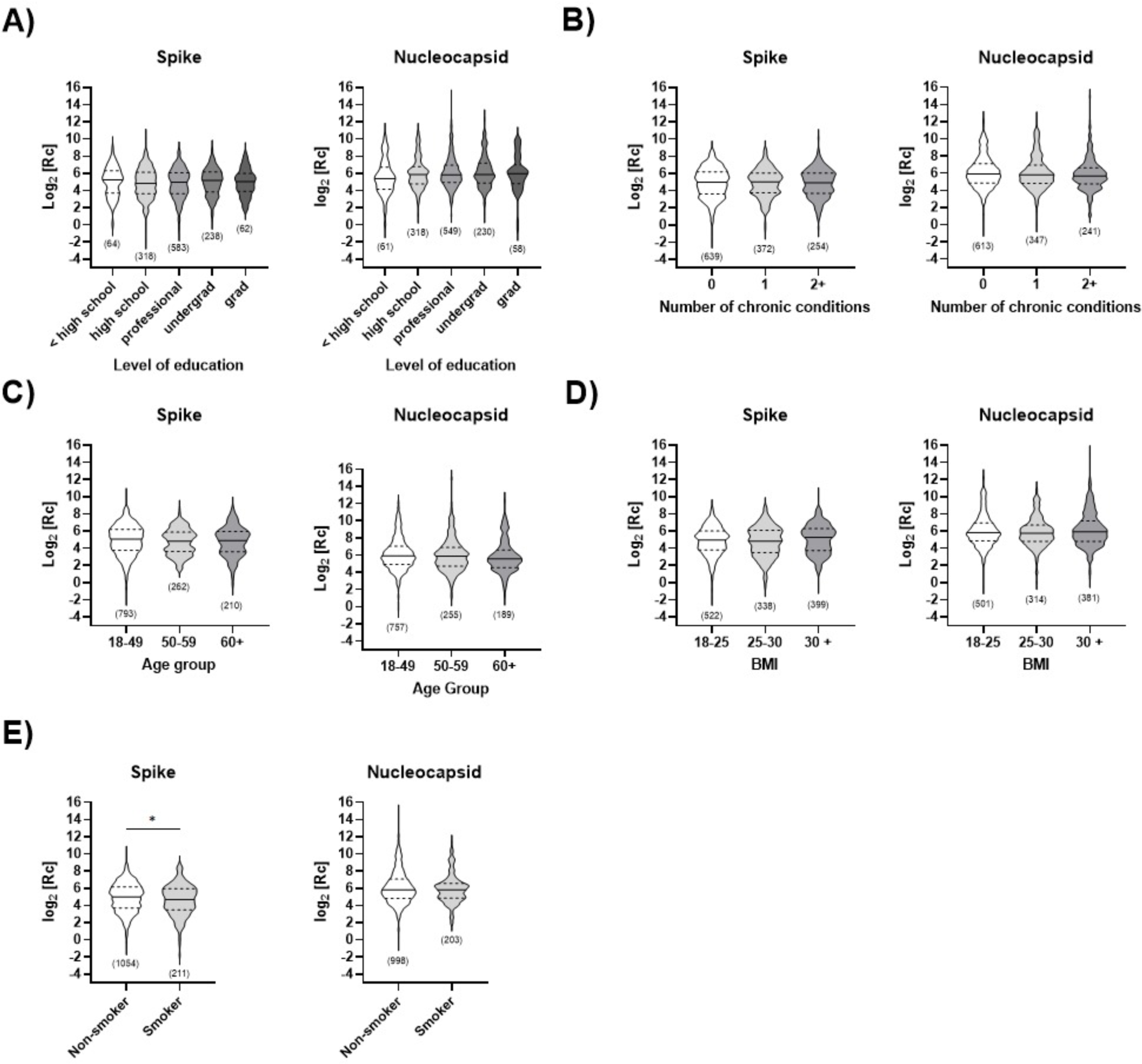
Impact of non-occupational activities on humoral immune response. ELISA measuring ancestral anti-spike or anti-nucleocapsid IgG levels are analyzed collectively (all visits together) and separated by **A)** level of education, **B)** number of chronic illnesses, **C)** age, **D)** BMI and **E)** smoking status. Results are shown in logarithmic form, post-transformation. The median (solid line) and quartiles (dashed lines) are shown in the violin plots; no outliers were removed. The total number of data points (n) per plot is shown. Statistical significance: *, p < 0.05. In **(D)**, two data points were removed because the category (BMI < 18) was not sufficiently populated to allow statistical analysis.

No statistically significant differences were found for any of the factors analyzed, except a statistically higher ancestral anti-spike IgG response in non-smokers than in smokers (p < 0.05). This difference was not observed in the case of natural immunity (anti-nucleocapsid IgG). This is consistent with prior observations that smokers generally show lower antibody titers or a more rapid decline of vaccine-induced IgG levels than non-smokers [59]. The results shown above were analyzed collectively (all visits together); no significant difference was observed when statistical analysis was stratified by visit.

## Discussion

This 1.5-year longitudinal study of the humoral response elicited by vaccination and by SARS-CoV-2 infection in a cohort of 304 essential workers from food service and retail in the Québec City area continues to highlight the capacity for a decentralized approach to effectively monitor the humoral response to infection, particularly in specific, understudied populations [37]. The impact of this study is amplified because cohort studies of highly vaccinated adults without pre-existing severe health problems who were not hospitalized for COVID-19 are scarce [40,41] and because few COVID-19 studies have reported longitudinal data from sampling of the same individuals over a long period [42].

We demonstrated data analysis first with the standard YES/NO determination of SARS-CoV-2 vaccine seroconversion and infection events. This confirmed high rates of seroconversion concomitant with each subsequent vaccination campaign and allowed to clearly observe seroconversion resulting from infection in this population of essential workers. We then obtained key information by quantifying the extent of humoral response. This required that the flexible in-house (decentralized) colorimetric ELISA protocol be tuned to minimize overly high absorbance datapoints that cannot be interpreted. To enable robust statistical analysis using Welch’s ANOVA test that requires normally distributed datasets, we applied data transformation to achieve the normal distribution; again, tuning the ELISA protocol was essential to produce unbiased transformed data.

In conclusion, the adaptability of the in-house colorimetric ELISA allowed rapid analytical intake of the new SARS-CoV-2 variant antigens as they became relevant to this population. By those means, we verified strong cross-reactivity between the ancestral and delta spike antigens, and slightly weaker cross-reactivity with the Omicron spike antigen. The results of ANOVA-Welsh statistical analysis are concordant with statistical differences reported for more widely studied cohorts such as healthcare workers, in the number of events of exposure to the antigen (vaccination or infection), in the effectiveness of vaccine types, and smoking. We further revealed differences related to occupational sectors over the period of the two first visits, during which the cohort mostly became fully vaccinated but where the rate of infection-acquired immunity was low. The flexibility of the decentralized analysis in rapidly integrating new iterations of the spike antigen and the ease of undertaking measurements in small laboratories at decentralized test sites will make this an essential complement to centralized testing in future epidemic or pandemic events.

## Data Availability

All data produced in the present study are available upon reasonable request to the authors.

## Acknowledgments

We acknowledge Yan Watts for his help with the validation of statistical analysis and Christian Gervais for his role in providing SARS-CoV-2 antigenic proteins.

## Supporting information captions

### Supporting Tables

**Table S1.** Number of data points removed for each antigen dataset.

**Table S2.** Percentage of fully vaccinated (at least two doses) individuals per visit.

### Supporting Figures

**Figure S1.** Seropositivity determined with in-house ELISA over 18 months.

**Figure S2.** Seropositivity determined with centralized ELISA over 18 months.

**Figure S3.** Overview of the ELISA IgM results over 18 months.

**Figure S4.** Impact of public transport use on immune response.

**Figure S5.** Impact of being in contact with a COVID-19 positive person on immune response.

**Figure S6.** Impact of BMI on immune response.

**Figure S7.** Comparative assessment of humoral immune response in unvaccinated and vaccinated individuals.

**Figure S8.** Comparative assessment of humoral immune response for different vaccine types.

