## Supporting Tables and Figures for "Longitudinal Study on Seroprevalence and Immune Response to SARS-CoV-2 in a Population of Food and Retail Workers Through Transformation of ELISA Datasets"

[1]Département de Chimie, Université de Montréal, Montréal, QC, Canada ; [2]PROTEO, Regroupement Québécois de Recherche sur la Fonction, l'Ingénierie et les Applications des Protéines, Québec, QC, Canada ; [3]Centre en Chimie Verte et Catalyse, Université de Montréal, Montréal, QC, Canada; [4]Départment de Biochimie et Médecine Moléculaire, Université de Montréal, Montréal, QC, Canada; [5]Mammalian Cell Expression, Human Health Therapeutics Research Centre, National Research Council Canada, Montréal, QC, Canada; [6]Centre de Recherche du Centre Hospitalier Universitaire de Québec, Université Laval, Québec, QC, Canada; [7]Département de Microbiologie-Infectiologie et d'Immunologie, Université Laval, Québec, QC, Canada; [8]Départment de Chimie, Université Laval, Québec, QC, Canada; [9]Centre d'Optique, Photonique et Laser, Université Laval, Québec, QC, Canada; [10]Institut Courtois, Université de Montréal, QC, Canada; [1111]Centre Québécois sur les Matériaux Fonctionnels, Regroupement québécois sur les matériaux de pointe, Centre Interdisciplinaire de Recherche sur le Cerveau et l'Apprentissage, Montréal, QC, Canada; [12]Department of Biochemistry, Microbiology and Immunology, Faculty of Medicine, University of Ottawa, Ottawa, ON, Canada; [13]Ottawa Center for Infection, Immunity and Inflammation (CI3), Ottawa, ON, Canada; [14] Département de Chimie, Université du Québec à Montréal, Montréal, QC, Canada

† These authors contributed equally

### Table of contents

**Supporting Tables**

**Table S1.** Number of data points removed for each antigen dataset.

**Table S2.** Percentage of fully vaccinated (at least two doses) individuals per visit.

**Supporting Figures**

**Figure S1.** Seropositivity determined with in-house ELISA over 18 months.

**Figure S2.** Seropositivity determined with centralized ELISA over 18 months.

**Figure S3.** Overview of the ELISA IgM results over 18 months.

**Figure S4.** Impact of public transport use on immune response.

**Figure S5.** Impact of being in contact with a COVID-19 positive person on immune response.

**Figure S6.** Impact of BMI on immune response.

**Figure S7.** Comparative assessment of humoral immune response in unvaccinated and vaccinated individuals.

**Figure S8.** Comparative assessment of humoral immune response for different vaccine types.

**Table S1. Number of data points removed for each antigen dataset.**

| | Ancestral spike | | Nucleocapsid | | Delta spike | | Omicron spike | |
|---|---|---|---|---|---|---|---|---|
| Visit | Data points removed | Total data points | Data points removed | Total data points | Data points removed | Total data points | Data points removed | Total data points |
| V1 | 1 | 303 | 12 | 303 | 56 | 304 | 6 | 304 |
| V2 | 7 | 297 | 34 | 294 | 19 | 297 | 36 | 297 |
| V3 | 2 | 291 | 12 | 292 | 3 | 289 | 1 | 290 |
| V4 | 0 | 194 | 2 | 192 | - | - | 0 | 194 |
| V5 | 1 | 191 | 2 | 182 | - | - | 0 | 194 |

**Table S2. Percentage of fully vaccinated (at least two doses) individuals per visit.**

| Visit | Participants having received at least 2 vaccine doses | Participants having completed the visit | Fully vaccinated participants (%) |
|---|---|---|---|
| V1 | 146 | 304 | 48 |
| V2 | 182 | 297 | 61 |
| V3 | 188 | 291 | 65 |
| V4 | 188 | 198 | 95 |
| V5 | 188 | 194 | 97 |

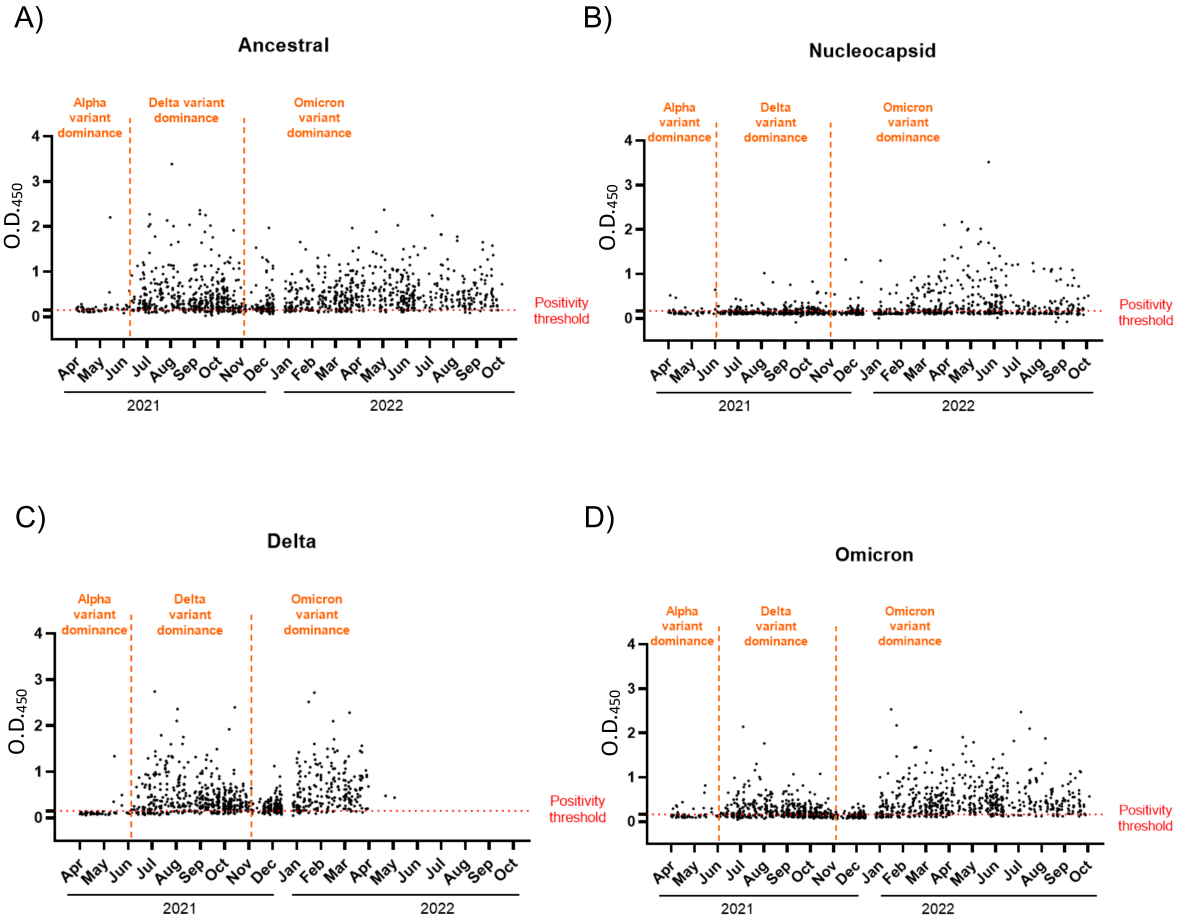


**Figure S1. Seropositivity determined with in-house ELISA over 18 months**. Determination of IgG for the **A)** ancestral spike, **B)** nucleocapsid, **C)** delta spike and **D)** Omicron antigens were measured. On the x-axis: the month of reference. On the y axis: $O.D._{450}$ for each sample measured.

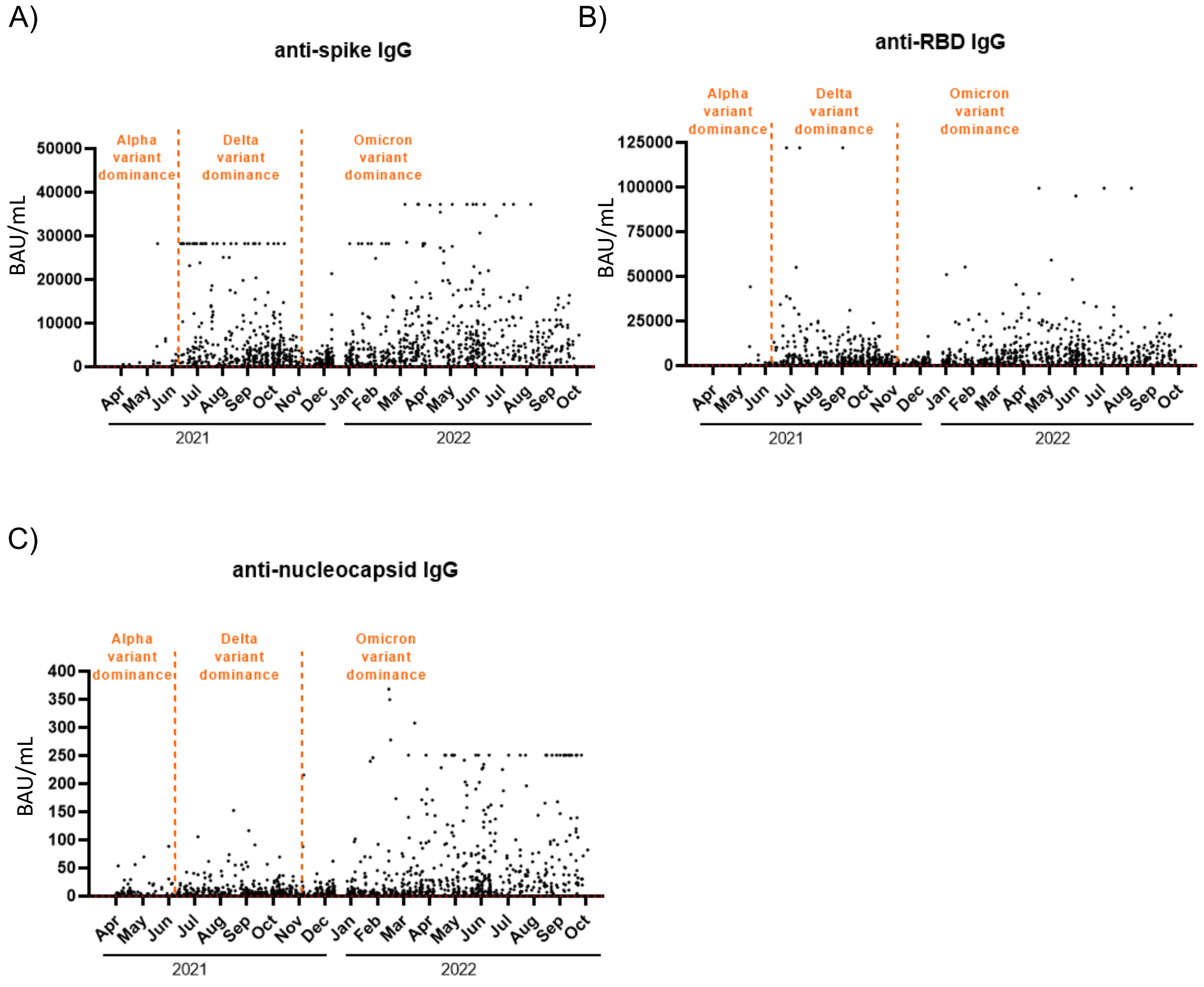


**Figure S2. Seropositivity determined with centralized ELISA over 18 months**. Determination of IgG for the **A)** ancestral spike, **B)** receptor binding domain (RBD) and **C)** nucleocapsid antigens were measured. On the x-axis: the month of reference. On the y axis: the concentration in BAU/mL for each sample measured.

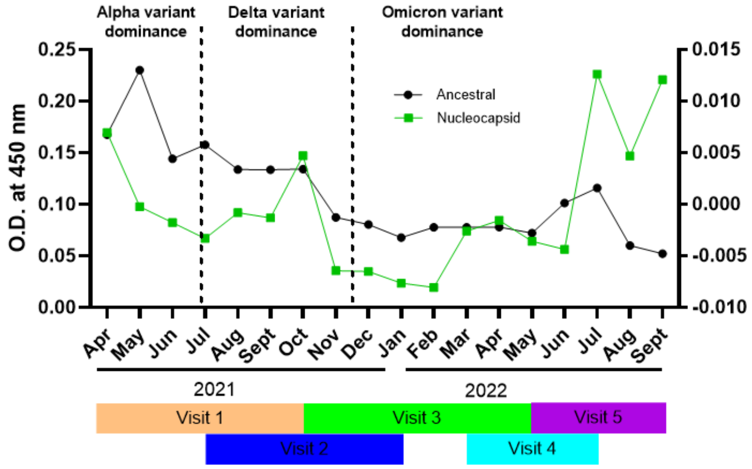


**Figure S3. Overview of the ELISA IgM results over 18 months.** Sample collection visits are shown in different colors under the graph. IgMs for the ancestral spike ectodomain and the nucleocapsid antigen were measured (see legend). On the x-axis: the month of reference. On the y axes: the mean $O.D._{450}$ of all samples collected during each given month. Note the different scale on the y-axes.

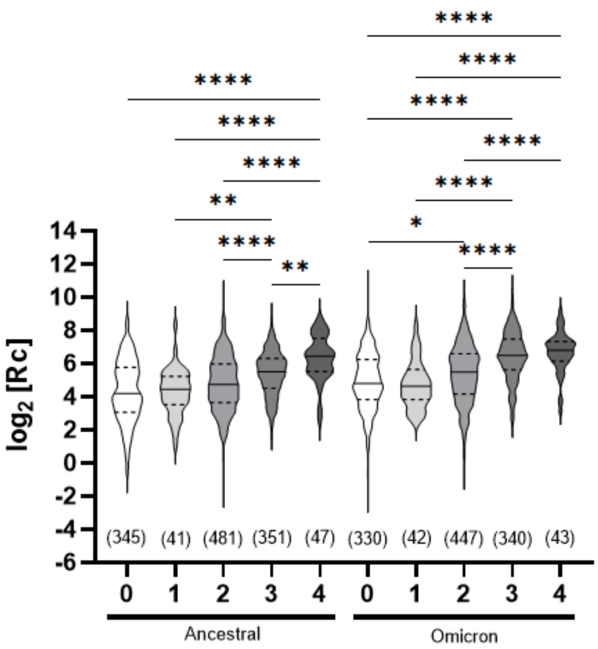


**Figure S4. Comparative assessment of humoral immune response in unvaccinated and vaccinated individuals including of individuals following seroconversion for anti-nucleocapsid IgG**. ELISA assays measuring ancestral or Omicron anti-spike IgG levels were analyzed collectively. X-axis: number of vaccine doses received (minimum 7 days post-vaccination). Each grouping includes all data collected (n is given) for that number of vaccine doses, including any data points taken at different visits for the same individual whose vaccination status did not change. For this reason, the number of datapoints may exceed the total number of participants (n = 304). Y-axis: data points after logarithmic transformation. The median (solid line) and quartiles (dashed lines) are shown in the violin plots; no outliers were removed. Statistical significance: *, $p < 0.05$; **, $p < 0.01$; ****, $p < 0.0001$.

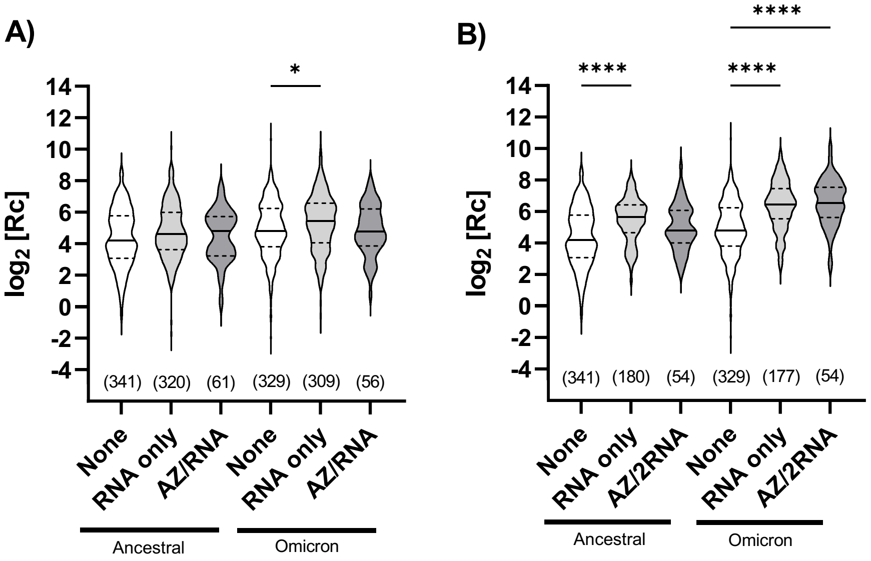


**Figure S5. Comparative assessment of humoral immune response for different vaccine types including of individuals following seroconversion for anti-nucleocapsid IgG**. ELISA assays measuring ancestral or Omicron anti-spike IgG levels were analyzed collectively. Samples following administration of **A)** 2 vaccine doses or **B)** 3 vaccine doses. "RNA only": either the Pfizer-BioNTech Comirnaty® mRNA COVID-19 vaccine, the Moderna Spikevax® mRNA COVID-19 vaccine, or a combination of both. "AZ/RNA" and "AZ/2RNA": one dose of the AstraZeneca COVISHIELD® viral vector-based COVID-19 vaccine and one or two doses of an RNA vaccine, respectively. Each grouping includes all data collected (n is given) for that type of vaccine, including any data points taken at different visits for the same individual whose vaccination status did not change. For this reason, the number of datapoints may exceed the total number of participants (n = 304). Individuals who had received the AstraZeneca vaccine only, or two doses of AstraZeneca and one of an RNA vaccine, were excluded as there was insufficient data to allow statistical analysis. Y-axes: data points after logarithmic transformation. The median (solid line) and quartiles (dashed lines) are shown in the violin plots; no outliers were removed. Statistical significance: *, $p < 0.05$; ****, $p < 0.0001$.

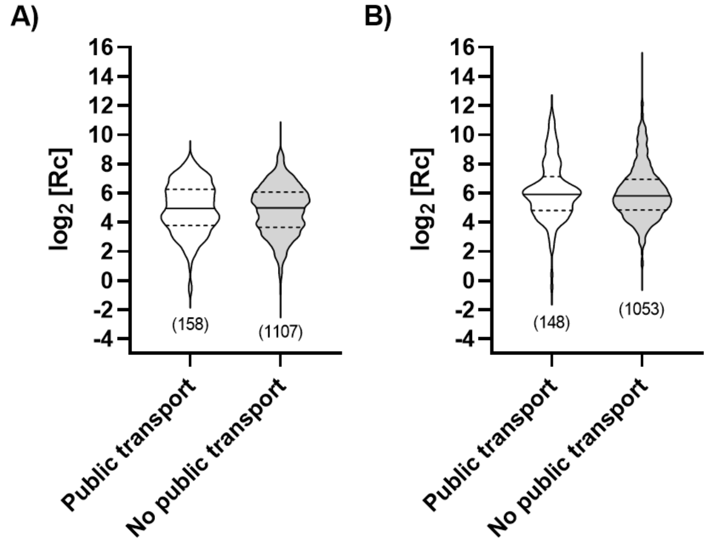


**Figure S6. Impact of public transport use on immune response**. ELISA measuring IgG for the **A)** ancestral variant spike ectodomain and **B)** nucleocapsid protein performed from visits 1 through 5 are analyzed together. Results are shown in post-transformation logarithmic form. The median (solid line) and quartiles (dashed lines) are shown in the violin plot where all values are included (no outliers removed). The number of data points (n) is shown in brackets below each dataset.

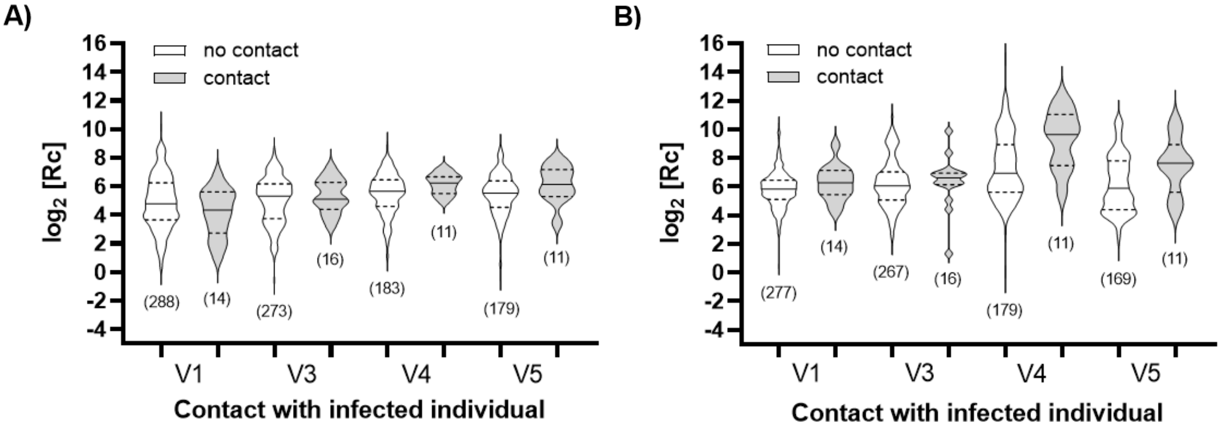


**Figure S7. Impact of being in contact with a COVID-19 positive person on immune response**. ELISA measuring IgG for the **A)** ancestral variant spike ectodomain and **B)** nucleocapsid protein performed from visits 1 through 5; visit 2 is not shown as there are not enough datapoints for statistical analysis. Results are shown in post-transformation logarithmic form. The median (solid line) and quartiles (dashed lines) are shown in the violin plot. The number of data points (n) is shown in brackets below each dataset.

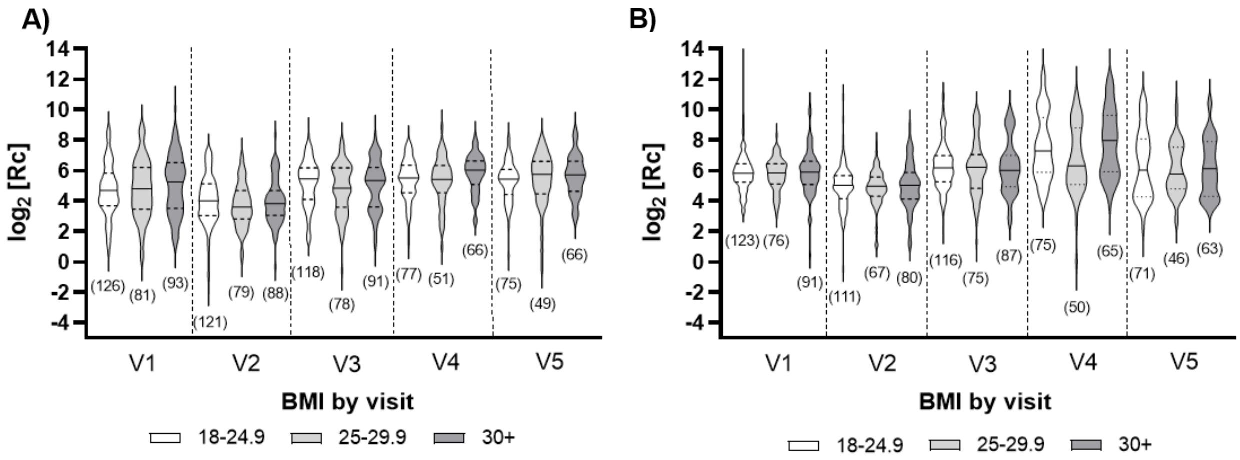


**Figure S8. Impact of BMI on immune response**. ELISA measuring IgG for the **A)** ancestral variant spike ectodomain and **B)** nucleocapsid protein performed from visits 1 through 5. Results are shown in post-transformation logarithmic form. The median (solid line) and quartiles (dashed lines) are shown in the violin plot. The number of data points (n) is shown in brackets below each dataset. Two datapoints (BMI < 18) were removed as the category did not have enough datapoints to allow statistical analysis.
